# Prevention of cardiac mortality through antihypertensive therapy using a combination of angiotensin-converting enzyme inhibitors and beta-blockers: a retrospective cohort study in Tsunan, Japan

**DOI:** 10.1101/2025.06.29.25330526

**Authors:** Shin-ichiro Ishikawa, Yusaku Hayashi, Toshiyuki Abe, Susumu Tanaka

## Abstract

Antihypertensive treatment significantly reduces the risk of cardiovascular mortality in patients with hypertension; however, treatment effects are weaker on cardiac mortality than on cerebrovascular mortality, even with equivalent blood pressure reduction. A potential cause of this difference is the activation of neurohumoral factors; however, few studies have addressed this gap. Therefore, in this study, we evaluated the effectiveness of a combination therapy prioritizing angiotensin-converting enzyme inhibitors and beta-blockers in reducing cardiac mortality in outpatients with hypertension. The retrospective observational cohort study spanned 30 years (1987–2016) at a single institution. From 1992 to 2001, we prioritized the use of a combination of angiotensin-converting enzyme inhibitors and beta-blockers to suppress neurohumoral factors, with calcium channel blockers and diuretics used as supplementary treatments. We tracked the standardized mortality ratios for all-cause, cardiac, and cerebrovascular mortality during each period. Using national data, we retrospectively compared the trends in all-cause and cardiovascular mortality rates in our region with those of the Japanese population. The standardized mortality ratios for all-cause and cardiac mortality in our region decreased significantly from 1992 to 2016 (that is, after treatment initiation) and were significantly lower than national averages after the initial control period. These findings suggest that antihypertensive therapy prioritizing the combination of angiotensin-converting enzyme inhibitors and beta-blockers may contribute to a reduction in cardiac mortality. Future prospective, randomized, double-blind controlled trials are crucial for validating this hypothesis.

## Introduction

The primary goal of hypertension treatment is to reduce overall mortality, as stated in the guidelines of the Japanese Society of Hypertension [1]. The current treatment approach for uncomplicated hypertension emphasizes blood pressure reduction, regardless of the antihypertensive drug used [2]. Strict blood pressure control reduces all-cause mortality and cardiovascular mortality significantly and is recommended in many guidelines [1,3]. However, some meta-analyses have suggested that the reduction in all-cause mortality through strict blood pressure is primarily due to a decrease in cerebrovascular mortality, with no significant effect on cardiac mortality [4]. Furthermore, a meta-analysis has indicated that strict blood pressure control does not reduce all-cause mortality or cardiovascular mortality [5].

Comparative trials of antihypertensive drugs with equivalent blood pressure reductions have shown significant differences in all-cause and cardiovascular mortality [6], and a meta-analysis has suggested that angiotensin-converting enzyme (ACE) inhibitors reduce all-cause and cardiac mortality [7]. This phenomenon is referred to as the “beyond blood pressure– lowering effect,” which suggests that factors other than blood pressure reduction, such as the suppression of neurohumoral factors, may be involved.

Observational studies have shown that cardiac mortality, especially fatal myocardial infarction and sudden cardiac mortality, is most frequent in the early morning, with activation of neurohumoral factors in the early morning contributing to this phenomenon [8–10]. ACE inhibitors suppress the renin–angiotensin system, which may explain their protective effect against cardiac mortality. However, the effects of beta-blockers remain debated [11,12]. The choice of a combination drug is also crucial. Diuretics and calcium channel blockers may activate the sympathetic nervous and renin–angiotensin systems, potentially weakening the effects of ACE inhibitors and beta-blockers [13–16]. Meta-analyses of interventional studies of patients with heart failure have shown that significant prognostic improvement occurs only when beta-blockers are combined with ACE inhibitors [17,18]. Interventional studies of heart failure have demonstrated the prognostic value of their combination. However, there are no reports of studies looking at the prognostic outcomes of their combination in hypertension.

We evaluated the effectiveness of a combination of ACE inhibitors and beta-blockers in reducing cardiac mortality in patients with hypertension. We hypothesized that suppressing neurohumoral factors, along with reducing blood pressure, is crucial for preventing cardiac mortality in patients with hypertension.

Accordingly, we prioritized combination therapy with ACE inhibitors and beta-blockers, while adding calcium channel blockers or diuretics as supplementary treatments when necessary. The impact of this treatment strategy on all-cause and cardiac mortality was investigated through a retrospective observational study.

## Materials and methods

### Study design and participants

This retrospective cohort study focused on all residents of Tsunan, Niigata, Japan, with a population of approximately 10,000. Tsunan is an agricultural town located in a mountainous area. The town’s population decreased from 12,941 in 1990 to 10,029 in 2015, whereas the proportion of residents aged 65 years and older increased from 23.0% to 39.0% [19]. The town has limited medical facilities, comprising only Tsunan Town Hospital and two clinics, with primary care for acute heart disease provided by the hospital. The study included approximately 2,000 patients with hypertension, representing 17% of the town’s population, who visited Tsunan Town Hospital from 1987 to 2016. All individual data were collected and analyzed by the Ministry of Health and Labor. We had access to the data, which are publicly available, from 15/11/2015 to 20/09/2020 for research purposes. The data used in this study were accessed for research purposes from 15/03/2021 to 22/05/2021. The researchers did not know the details for each individual, including the length of hospitalization.

A treatment strategy prioritizing the use of ACE inhibitors and beta-blockers to suppress neurohumoral factors was implemented, with calcium channel blockers and antihypertensive diuretics added as needed. This approach was termed the “ABCD strategy” (A = ACE inhibitors, B = beta-blockers, C = calcium channel blockers, and D = diuretics). This strategy was implemented from 1992 to 2006, after which treatment decisions were left to the discretion of attending physicians. The 5-year period from 1987 to 1991, before the implementation of this strategy, was defined as the control period. The entire observation period was divided into six 5-year intervals: Period 0 (1987–1991), Period 1 (1992–1996), Period 2 (1997–2001), Period 3 (2002–2006), Period 4 (2007–2011), and Period 5 (2012–2016). Using Period 0 as the baseline, trends in mortality-related indicators during the subsequent periods were retrospectively analyzed and compared with nationwide data for Japan.

### Treatment protocol

The participants were outpatients with hypertension, including all new patients with a morning home systolic blood pressure of 135 mmHg or higher. Patients receiving other antihypertensive drugs were switched to a combination of ACE inhibitors and beta-blockers whenever possible. Patients were excluded if they had decompensated heart failure, symptomatic bradycardia (resting heart rate <50 beats/min without pacemaker treatment), acute myocardial infarction or stroke, an increase in creatinine levels by 30% or more or by 1 mg/dL or more from pre-administration levels after 1 month of medication, critical side effects such as liver dysfunction, or decreased quality of life. Patients were also excluded if they were deemed inappropriate for participation in the study by the attending physician.

The ACE inhibitors used were enalapril [7], trandolapril [20], and perindopril [7], which have been shown to reduce the number of fatal events in several interventional trials. The maximum maintenance doses were 10, 2, and 8 mg/d, respectively.

Similarly, beta-blockers associated with reduced mortality, such as metoprolol [21], carvedilol [22], and bisoprolol [23], were used. For beta-blockers, in patients with a history of heart failure or suspected latent cardiac dysfunction with a brain natriuretic peptide level higher than 100 pg/dL, treatment was started at one-fourth of the usual dose. If no new-onset or exacerbation of heart failure or symptomatic bradycardia occurred, the dose was doubled every month to the maximum maintenance doses of 120, 20, and 5 mg/d, respectively.

Among calcium channel blockers, the long-acting dihydropyridine amlodipine was selected because short-acting calcium channel blockers are associated with a risk of cardiac mortality [24]. For antihypertensive diuretics, if the pre-administration serum potassium level was <4.5 mEq/L, spironolactone (50 mg/d), an aldosterone antagonist used in the RALES trial [25], was used. In cases of gynecomastia, spironolactone was replaced with eplerenone (50 mg/d) [26]. For serum potassium levels >4.5 mEq/L, indapamide (2 mg/d), a non-thiazide diuretic, was used. Indapamide has been reported to be associated with better cardiovascular outcomes than those of thiazide diuretics [27].

Since the late 1990s, angiotensin II receptor antagonists have been widely used in Japan. However, as their effect on preventing cardiac mortality was unclear [28], they were excluded from the drug selection during this period, unless there was a special reason.

The target for blood pressure reduction was a morning home systolic blood pressure lower than 135 mmHg. Based on reports that combination therapy has advantages even in mild hypertension [29], treatment was initiated with a combination of low-dose ACE inhibitors and beta-blockers. For patients already undergoing antihypertensive treatment, existing antihypertensive drugs were reduced while switching to ACE inhibitors and beta-blockers, ensuring that the morning home systolic blood pressure did not exceed 135 mmHg.

If the blood pressure target was not achieved with these combinations, calcium channel blockers and antihypertensive diuretics were added to the treatment regimen. This selection order was based on the results of the ACCOMPLISH trial [30], which showed that calcium channel blockers are significantly more effective than antihypertensive diuretics in reducing cardiovascular mortality when combined with ACE inhibitors.

Antihypertensive drugs were primarily administered at bedtime. Although a recent report suggests that the timing of administration does not affect cardiovascular mortality [31], in the present study, drugs were administered before bedtime based on a report indicating that this timing has a greater antihypertensive effect and reduces cardiovascular mortality [32].

If the target blood pressure could not be achieved even with the administration of four drugs, other antihypertensive drugs were added at the discretion of the attending physician. The achievement rate of the blood pressure target was calculated from medical record entries for each period every 5 years.

### Ethical considerations

The data used in this study were exempt from review by the Ethics Review Committee of the Jikei University School of Medicine [receipt number 34-413 (11570)]. The selection of therapeutic agent was discussed with participants. Upon selection of the drug, consent was obtained. All data are processed and stored by the MHLW and the public health centers and were already anonymized. Individual data, including names, or data that would identify individuals, are not available. Therefore, the Ethics Committee approved the use of the data without the need to obtain informed consent from each individual, and we have received a waiver of this requirement.

### Clinical outcomes and endpoints

To evaluate the results of this study, cause-specific mortality statistics, which are highly objective and regularly reported by the Ministry of Health, Labour and Welfare as well as local municipal health and welfare departments, were used [33]. The primary endpoints were all-cause mortality and cardiovascular mortality every 5 years. The secondary endpoint was death due to a malignant neoplasm. The standardized mortality ratio (SMR) was used as the evaluation index. This measure is used to eliminate bias due to differences in age composition among small population-scale municipalities [34].

### Statistical analyses

The SMR and its 95% confidence interval (CI) were calculated and compared with the national average of 1.0 [34]. The SMR was calculated using the Niigata Prefecture Medical Representatives Calculation Sheet (version 0.71) created by Susumu Tanaka. The total population was used for the population figures. The proportion of antihypertensive drug use was compared with national data based on drug prices, using the average frequency of antihypertensive drug use each year for Periods 0 (1991), 2 (1998), and 5 (2014). The number of antihypertensive drugs used at the hospital was calculated from the total annual price of all antihypertensive drugs prescribed during the relevant year and stored in the pharmacy. National data were obtained from Intercontinental Marketing Services Health Inc. (IMS) [35], IQVIA (I [IMS Health], Q [Quintiles], and VIA [by way of]) [36], and National Database Open Data [37] with permission.

## Results

### Trends in blood pressure target achievement rates

Approximately 2,000 patients with hypertension were treated using the ABCD strategy. The blood pressure target achievement rates based on outpatient blood pressure measurements of 500 randomly selected patients with hypertension was approximately 50% in Period 0. In Periods 1 and 2, based on early-morning home blood pressure measurements from 500 patients with hypertension, the achievement rates for the target blood pressure with the two-drug combination therapy were 50% and 52%, respectively, and when patients receiving up to four drug combinations were included, the rates were 63% and 65%, respectively. In Periods 3, 4, and 5, which included patients receiving up to four drug combinations, the achievement rates remained greater than 60% (63%, 67%, and 61%, respectively).

### Trends in antihypertensive drug use

Nationwide, the combined use of ACE inhibitors and beta-blockers decreased from 41.4% in Period 0 to 38.5% in Period 2 and 14.6% in Period 5 (Table 1). Conversely, in Tsunan Town Hospital, usage rates increased from 40.1% in Period 0 to 64.6% in Period 2 but decreased to 47.6% in Period 5.

**Table 1.** Drug Use Proportions in Japan and Tsunan Hospital Over Time.

|  | Year | ACEI (%) | BB (%) | CCB (%) | ARB (%) | Others (%) |
| --- | --- | --- | --- | --- | --- | --- |
| **Japan** | **1991** | 23.9 | 17.5 | 45.8 | 12.7 | 0 |
|  | **1998** | 24.8 | 13.7 | 50.7 | 10.1 | 0.7 |
|  | **2014** | 7.3 | 7.3 | 20.1 | 64 | 1.3 |
| **Tsunan Hospital** | **1991** | 10.1 | 30 | 51.6 | 8.3 | 0 |
|  | **1998** | 26.3 | 38.3 | 16.9 | 17.3 | 0.9 |
|  | **2014** | 25.9 | 21.7 | 22 | 16.8 | 13.6 |

ACEI: angiotensin-converting enzyme inhibitors, BB: beta-blockers, CCB: calcium channel blockers, ARB: angiotensin II receptor blockers, Others: other medications.

Nationally, the use of calcium channel blockers increased from 45.8% in Period 0 to 50.7% in Period 2 but decreased to 20.1% in Period 5 because of the increased use of angiotensin II receptor antagonists. In Tsunan Town Hospital, the use of calcium channel blockers decreased from 51.6% in Period 0 to 16.9% in Period 2 and then increased to 22.0% in Period 5.

### Trends in SMRs in Tsunan

Table 2 shows the trends in the SMRs (95% CI) for all-cause mortality and deaths due to heart disease, cerebrovascular disease, and malignant neoplasms from Periods 0 to 5. The SMRs for all-cause mortality in Tsunan in Periods 0, 1, 2, 3, 4, and 5 were 0.94, 0.88, 0.80, 0.87, 0.78, and 0.84, respectively, whereas those for cardiac mortality were 0.91, 0.77, 0.55, 0.73, 0.72, and 0.81, respectively.

**Table 2.** Standardized Mortality Ratios and 95% Confidence Intervals for All Deaths and the Three Major Causes of Death in Japan and Tsunan Town Over Time.

|  | **Period 0** | **Period 1** | **Period 2** | **Period 3** | **Period 4** | **Period 5** |
| --- | --- | --- | --- | --- | --- | --- |
|  | **1987–1991** | **1992–1996** | **1997–2001** | **2002–2006** | **2007–2011** | **2012–2016** |
| **All deaths** |  |  |  |  |  |  |
| Japan | 1.00 | 1.00 | 1.00 | 1.00 | 1.00 | 1.00 |
| Tsunan Town |  |  |  |  |  |  |
| Total | 0.94 (0.88–1.01) | 0.88 (0.79–0.97) | 0.80 (0.74–0.86) | 0.87 (0.81–0.93) | 0.78 (0.73–0.84) | 0.84 (0.78–0.89) |
| Male | 0.93 (0.84–1.03) | 0.91 (0.83–1.00) | 0.82 (0.73–0.90) | 0.95 (0.83–1.04) | 0.86 (0.77–0.94) | 0.88 (0.80–0.96) |
| Female | 0.95 (0.85–1.05) | 0.88 (0.79–0.97) | 0.78 (0.69–0.86) | 0.78 (0.70–0.86) | 0.72 (0.65–0.79) | 0.80 (0.73–0.87) |
| **Malignant neoplasm** |  |  |  |  |  |  |
| Japan | 1.00 | 1.00 | 1.00 | 1.00 | 1.00 | 1.00 |
| Tsunan Town |  |  |  |  |  |  |
| Total | 0.86 (0.73–1.00) | 0.78 (0.69–0.80) | 0.68 (0.57–0.78) | 0.87 (0.76–0.99) | 0.84 (0.73–0.95) | 0.95 (0.83–1.07) |
| Male | 0.86 (0.70–1.03) | 0.94 (0.81–1.07) | 0.69 (0.56–0.83) | 0.96 (0.80–1.12) | 0.92 (0.77–1.08) | 0.92 (0.76–1.08) |
| Female | 0.86 (0.65–1.07) | 0.83 (0.68–0.97) | 0.64 (0.48–0.80) | 0.75 (0.59–0.92) | 0.73 (0.57–0.88) | 0.99 (0.80–1.17) |
| **Heart disease** |  |  |  |  |  |  |
| Japan | 1.00 | 1.00 | 1.00 | 1.00 | 1.00 | 1.00 |
| Tsunan Town |  |  |  |  |  |  |
| Total | 0.91 (0.77–1.05) | 0.77 (0.57–0.97) | 0.55 (0.43–0.68) | 0.73 (0.60–0.87) | 0.72 (0.60–0.84) | 0.81 (0.68–0.94) |
| Male | 0.87 (0.67–1.07) | 0.88 (0.70–1.08) | 0.52 (0.35–0.69) | 0.76 (0.56–0.97) | 0.78 (0.58–0.99) | 0.87 (0.68–1.12) |
| Female | 0.94 (0.74–1.14) | 0.82 (0.64–0.99) | 0.58 (0.41–0.75) | 0.71 (0.53–0.88) | 0.68 (0.52–0.83) | 0.75 (0.59–0.91) |
| **Cerebrovascular disease** |  |  |  |  |  |  |
| Japan | 1.00 | 1.00 | 1.00 | 1.00 | 1.00 | 1.00 |
| Tsunan Town |  |  |  |  |  |  |
| Total | 1.08 (0.89–1.26) | 0.90 (0.66–1.13) | 0.88 (0.72–1.04) | 0.88 (0.72–1.05) | 0.88 (0.71–1.05) | 1.21 (0.99–1.40) |
| Male | 1.08 (0.81–1.35) | 0.97 (0.79–1.16) | 1.00 (0.75–1.25) | 1.16 (0.88–1.45) | 0.89 (0.62–1.15) | 1.14 (0.81–1.46) |
| Female | 1.07 (0.82–1.32) | 1.13 (0.92–1.33) | 0.77 (0.57–0.98) | 0.65 (0.46–0.85) | 0.88 (0.66–1.11) | 1.25 (0.97–1.54) |

Compared with that in Period 0, the SMRs for all-cause mortality and cardiac mortality decreased significantly after the introduction of the combination therapy. The SMR for cardiac mortality gradually increased after Period 3; however, it remained low (Fig 1).

**Fig 1.**
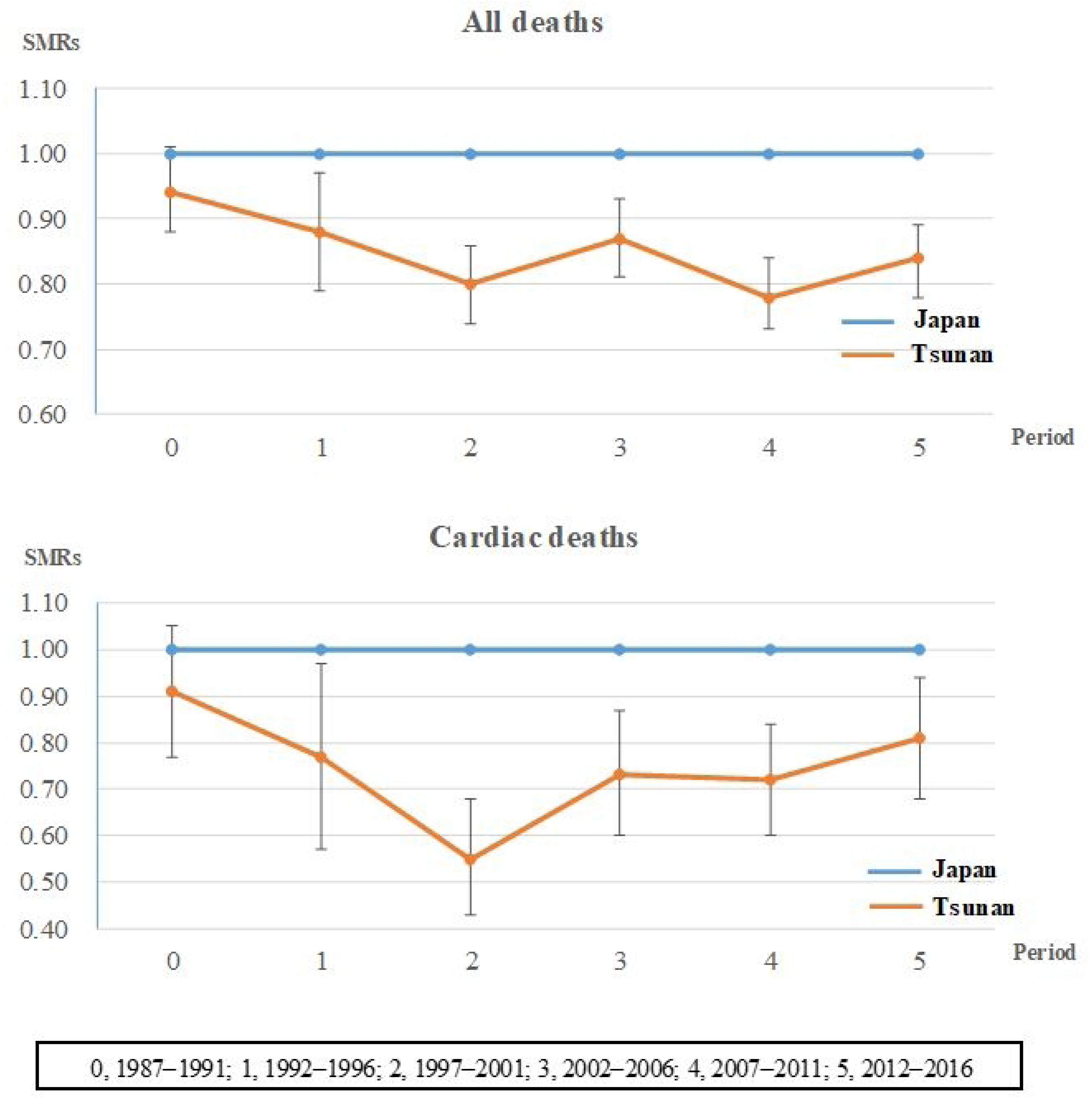
Standardized Mortality Ratios for All Deaths and Cardiac Mortality in Every 5-year Period.

## Discussion

In this study, we tested the hypothesis that neurohumoral factor activation influences the risk of all-cause mortality, especially cardiac mortality, following antihypertensive treatment. Compared with national data, the SMR for cardiac mortality in Tsunan showed no significant difference in Period 0 but began to decrease from Period 1 after changing the combination drug from calcium channel blockers to ACE inhibitors. This result suggests that combination therapy with ACE inhibitors and beta-blockers contributes to a decrease in mortality.

Although all-cause mortality in Tsunan showed no significant difference from the national average in Period 0, it decreased significantly throughout the subsequent periods. This result can be partly attributed to a reduction in cardiac mortality. Given that the blood pressure target achievement rate in Tsunan did not differ significantly from the national average [1], both the degree of blood pressure reduction and the choice of antihypertensive drug combination may have influenced the results. Moreover, the persistent effect of this combination therapy for over 20 years suggests the possibility of a legacy effect due to delayed cardiovascular remodeling.

This was a real-world cohort study, likely including many high-risk hypertension cases with secondary prevention of cardiac mortality and complications, such as diabetes and dyslipidemia, in which neurohumoral factors are prone to elevation. A combination of ACE inhibitors and beta-blockers may have been particularly effective in these patients.

In contrast, the SMR for cerebrovascular mortality was not significantly different from the national average throughout the study period. This suggests that cerebrovascular mortality depends more on blood pressure reduction than on the type of antihypertensive drug used, consistent with the trends observed in many previous intervention studies.

Intervention studies in patients with heart failure have shown that ACE inhibitors [38] and beta-blockers [17] contribute significantly to an improved prognosis and that their combination is important [18]. In contrast, intervention studies using calcium channel blockers or diuretics have not shown improvements in prognosis [39,40]. These differences may be due to differences in the activation of neurohumoral factors [41,42].

The results of this study highlight the importance of suppressing the activation of neurohumoral factors during hypertension treatment. In the future, interventional prospective comparative studies of groups that use antihypertensive drugs that suppress neurohumoral factors and those that do not, with matched degrees of blood pressure reduction and targeting cases at high risk for cardiac mortality, will be necessary.

The decrease in all-cause mortality observed in this study could also be related to a reduction in the number of deaths due to malignant neoplasms. There are numerous risk factors for death due to malignant neoplasms. Observational studies have reported that ACE inhibitors and beta-blockers tend to decrease deaths from malignant neoplasms, which calcium channel blockers and antihypertensive diuretics increase deaths [43]. The effects of treatments on malignant neoplasms and contribution of deaths due to malignant neoplasms to our findings are topics for future investigations.

This study had some limitations. We primarily evaluated data based on clinical diagnoses recorded on death certificates. Although the number of deaths is accurate, the cause of death may not be accurate because it was not based on autopsies. Confounding factors affecting mortality, such as the prevalence of comorbidities (e.g., heart failure, ischemic heart disease, diabetes, dyslipidemia, and smoking habits), should be considered along with hypertension. However, we were unable to obtain longitudinal data for these factors during the follow-up period. In this study, no other interventions that could reduce all-cause mortality or cardiac mortality were implemented; therefore, it is not likely that factors other than antihypertensive treatment significantly influenced the prognosis of heart disease. Additionally, we were unable to track trends for a small number of patients with hypertension who may have received treatment at medical institutions outside the primary medical care area, other than the two clinics in Tsunan.

## Conclusion

Interventional studies focused on heart failure have shown a significant reduction in cardiac death in response to combination therapy with ACE inhibitors and beta-blockers, suggesting the involvement of neurohumoral factor activation. This concept has not been considered in the treatment of hypertension, and analyses of the outcomes of this combination therapy are lacking. In the present study, all-cause mortality and cardiac mortality were reduced in patients receiving antihypertensive therapy combining both ACE inhibitors and beta-blockers. These findings suggest that activation of neurohumoral factors is involved in the occurrence of cardiac mortality in hypertensive patients.

Although this was not an intervention study, it was a real-world clinical cohort study, without strict selection criteria. Therefore, the results have broad external validity and should provide useful insights for general clinical practice. Furthermore, to our knowledge, there are no long-term follow-up studies of such combination therapy over 30 years. In the future, it would be worthwhile to conduct a randomized controlled trial using this strategy in hypertensive patients with concomitant cardiac disease in which neurohumoral factors are likely to be activated to further evaluate the role of these factors.

## Data Availability

All of the data used in this study are publicly available and referenced in the paper. Specifically: Population Census data are available from the e-Stat database (https://www.e-stat.go.jp/stat-search/files?page=1&layout=datalist&toukei=00200521&tstat=000001049104&cycle=0&tclass1=000001049105). The Annual Report on Health and Welfare Statistics is available from the Niigata Prefecture website (https://www.pref.niigata.lg.jp/sec/fukushihoken/nenpo-reiwa02-01.html).

## Acknowledgments

We express our deepest gratitude to the following members for their cooperation in conducting this study: M. Tsuchiya, A. Sasaki, T. Sakamoto, S. Nakata, H. Suzuki, T. Hanafusa, K. Miyoshi, Y. Tanaka, F. Okazaki, T. Yasuzawa, A. Matsuyama, M. Uemura, T. Ito, and J. Koga. We would also like to thank Editage (www.editage.jp) for English language editing.

## Notes

### Competing Interest Statement

In this study, the SMR values were calculated using the Niigata Prefecture SMR Calculation Sheet Version 0.71 devised by Susumu TANAKA. This report concerns an investigator-initiated study, and the authors have no conflicts of interest to disclose with respect to this report.

### Funding Statement

The author(s) received no specific funding for this work.

### Author Declarations

The data used in this study were exempt from review by the Ethics Review Committee of the Jikei University School of Medicine [receipt number 34-413 (11570)]. Participants were included in the study after the advantages and disadvantages of the antihypertensive medications were verbally explained to them and they provided informed consent.

